# Expression and relationship with immunity of LRFN4 in lung adenocarcinoma: Based on bioinformatics analysis

**DOI:** 10.1101/2022.11.04.22281961

**Authors:** Zhiyong Zhu, Linglong Peng, Haoyun Luo, Yahui Jiang, Mingying Yang, Haitao Gu, Yaxu Wang

## Abstract

Leucine rich repeat and fibronectin type III domain containing 4(LRFN4) has been reported to be upregulated in multiple tumors and related to prognosis and survival of patients. However, the function of LRFN4 in LUAD is still unclear. Herein, bioinformatic approach was used for the first time to elucidate the relationship between LRFN4 and LUAD. In LUAD tissues, we discovered that LRFN4 mRNA expression was considerably higher. Higher LRFN4 expression was associated with poorer prognosis and higher clinical stage of LUAD patients. Paraffin pathology sections (12 samples including LUAD tissues and paired normal tissues from the Second Affiliated Hospital of Chongqing Medical University) were used to verify the expression of LRFN4 at the protein level by immunohistochemical staining. On the other hand, we identified that LRFN4 expression was related to multiple immune cells that constitute tumor immune microenvironment. Pathway enrichment analysis also suggested the enrichment of several tumor- and immune-related pathways, such as: Hippo pathway, NOD-like pathway, TNF pathway and P53 pathway. Finally, we constructed an 8-gene prognostic risk signature based on 35 LRFN4-related immunomodulators using the Cox regression model, and obtained reasonably good accuracy through Receiver Operating Characteristic curve (ROC curve) validation. The risk signature was further identified as an independent risk factor – was linked with worse survival of LUAD patients. Furthermore, a prognostic risk profile based on LRFN4-related immunomodulators was constructed. Meanwhile, other clinical features were integrated together as prognostic markers to construct a nomogram to predict the long-term survival probability of LUAD patients, and fairly high credibility was obtained by validation of calibration curves.

## Introduction

For several decades, lung cancer has been the most common kind of cancer and the leading cause of cancer deaths worldwide, with around 1.8 million new cases and 1.6 million deaths every year.[1, 2]Lung cancer has a 5-year survival rate of roughly 18%, owing primarily to late detection and a lack of therapy prescription[3, 4]. Non-small cell lung cancer (NSCLC) and small cell lung cancer (SCLC) are two types of lung cancer[5]. NSCLC, which includes squamous cell lung carcinoma, large cell lung carcinoma, and lung adenocarcinoma (LUAD), accounts for roughly 85 percent of lung cancers[2, 6]. LUAD is the most frequent subtype among them, accounting for around 40% of diagnoses [7, 8].

Cancer immunotherapy is a developing type of therapy that has been developed in the last decade. The therapy works by training immune cells to remove target cells which carry tumor antigens and eliminating immunosuppressive signals on the other[9]. The tumor microenvironment (TME) is a dynamic and changing environment composed with tumor cells, immune cells, fibroblasts, extracellular matrix, cytokines and other molecules, such as immune checkpoint key molecules, all these components surround and support the tumor nest, which greatly influence tumor proliferation, invasion and metastasis[10]. With the recent advent of new therapies, immunotherapy has emerged as an invaluable treatment option for patients with lung cancer[11]. Programmed cell death protein 1 (PD-1) and PD-L1, have emerged as main targets for immunotherapy in lung cancer. However, many lung cancers have less frequently express PDL1, which underlie the lack of response to immunotherapy[12]. Therefore, new targets for immunotherapy are urgently needed.

The protein-coding gene LRFN4 (leucine rich repeat and fibronectin type III domain containing 4) belongs to the SALM family, a family of synaptic adhesion molecules containing five members, LRFN4 is the third one, so it is also known as SALM3[13]. Previous research has linked LRFN4 expression to the chance of dying from a variety of malignancies, including: gastric cancer, colorectal cancer and ovarian cancer[13-17]. However, LRFN4’s function in lung cancer is uncertain.

## Materials and Methods

### Data acquisition and processing

RNA-seq data (normalized by Fragments Per Kilobase Million) and clinical information (including ensemble ID, age, survival time, survival status, T, N, M, and pathological stage) of LUAD patients were downloaded from The Cancer Genome Atlas website (TCGA https://portal.gdc.cancer.gov/). A total of 551 samples were incorporated, including 497 cancer tissues and 54 para-cancerous tissues. Perl software (version 5.30.2.1) was used to process these raw data: converting ensemble ID to gene name, etc. and eventually the above date was obtained in a readable format.

### Expression and clinical relevance analysis of LRFN4

The differences in LRFN4 expression between LUAD and normal tissues were investigated by R software (version 4.0.5) with the “limma” package, and visualized with “ggplot2”, “ggpubr” packages. The relationship between the expression level of LRFN4 and clinicopathological stage was conducted with independent sample *t*-test. Kaplan-Meier analysis was conducted with the “survival” package and visualized with “survminer” package of R software. Statistical significance was determined by P< 0.05.

### Immunohistochemical staining

The study protocol was approved by the ethics review board of Chongqing Bishan District People’s Hospital (approval number 2022-KY-13). We have obtained written informed consent from all study participants. Patients’ name were recorded and corresponded to paraffin sections so that clinical information can be obtained later.12 paraffin sections (from 12 LUAD patients who underwent surgery at the Second Affiliated Hospital of Chongqing Medical University from January 28th, 2022 to April 26th, 2022) were randomly selected for immunohistochemical staining analysis. Paraffin sections were all prepared by the same experienced pathologist and the quality was examined by another pathologist. Immunohistochemical staining was performed by the following steps according to the antibody instructions: (i) Paraffin sections were dewaxed, and the activity of endogenous peroxidase was blocked with 0.3% hydrogen peroxide; (ii) Sections were incubated by an anti-LRFN4 primary antibody(Rabbit Anti-LRFN4 antibody bs-11089R 1:500) at 4°C overnight; (iii) Sections were incubated with secondary antibody (Sheep anti-rabbit antibody PV-9001 ready to use); (iv) The sections were counterstained with hematoxylin and visualized by DAB solution; (v) Stained images were acquired by microscopy (Olympus, BX43) at magnifications of x20 and x40.

### Immune cell infiltration analysis

CIBERSORT is currently the most frequently used immune cell infiltration analysis tool. It inversely infers the proportions of various cells and assigns the results circularly with the method of deconvolution[18]. In this study, we set the number of cycles to 1000, and the content of each type of immune cell in every sample was calculated by CIBERSORT for subsequent analysis. The “vioplot” package was used to plot the infiltration level of immune cell in normal and tumor tissues. TIMER database is a widely used database which provides many ways to analyze tumor immunity (http://timer.cistrome.org/). Relationship between LRFN4 copy number variation and several immune cell infiltration levels was analyzed by TIMER1.0 database. With the median LRFN4 content as cut-off value, all samples were separated into two groups: those with high LRFN4 expression and those with low LRFN4 expression. The contents of 22 immune cells were calculated separately, and t-test was conducted between them.

TISIDB website (http://cis.hku.hk/TISIDB/) was performed for analyzing the immune implications of LRFN4. It pre-computes associations between multiple genes and immune features, such as lymphocytes, immunomodulators and chemokines[19]. Relationship between LRFN4 and 28 immune cell types was analyzed sequentially in LUAD. Meanwhile, LRFN4-related immunomodulators were obtained, which were used for subsequent construction of PPI network and prognostic risk signature.

### Functional enrichment analysis

Co-expressed molecules of LRFN4 which were used for functional enrichment analysis were obtained from CCLE database (https://moodle2.sscnet.ucla.edu/) (Co-expression coefficient greater than 0.5 and P-value less than 0.05). Gene Ontology analysis (GO) and Kyoto Encyclopedia of Genes and Genomes (KEGG) analysis were performed in R software with “clusterProfiler”,”enrichplot” and “pathview” packages. GSEA software (version 4.0.3) was used to find more pathways related to LRNF4 expression. During the analysis, we set the number of permutations to 1000. PPI network was constructed on STRING website(https://cn.string-db.org/) using immunomodulators obtained from TISIDB website (medium confidence).

### Establishment of predictive model

Univariate COX analysis was used to screen for immune prognostic related genes that combine prognosis and LRFN4 related immunomodulators. Multivariate Cox regression analysis was used to construct an immune-related prognostic risk model eventually. Riskscore was generated for each LUAD patient. To split all samples into high risk and low risk groups, the median Riskscore was used as a cut-off number. Kaplan-Meier analysis was performed for risk model prognosis analysis. To further verify the independent prognostic significance of Riskscore, clinical features such as age, gender and stage were combined with Riskscore to perform univariate and multivariate COX analysis by R software “survival” package. ROC curves were produced to evaluate predictive accuracy of Riskscore and clinical characteristics by “timeROC” package. Later, a nomogram containing Riskscore, age, gender and pathological stage was constructed, and visualized calibration curves for 3-year and 5-year were conducted to test its accuracy by “rms” package.

## Results

### The mRNA expression of LRFN4 was upregulated and related to clinical outcomes in LUAD

551 samples from the TCGA database were used to examine the difference in LRFN4 mRNA expression levels between tumor and paraneoplastic tissues in LUAD. As shown in Figure 1A, mRNA expression of LRFN4 was significantly upregulated in LUAD samples (P <0.001). Immunohistochemical staining was performed to verify the difference in LRFN4 protein expression level between tumor and normal tissues. We performed Kaplan-Meier survival analysis to determine whether LRFN4 expression and clinical outcomes were correlated. In Figure 1B, the survival probability of high LRFN4 expression group was lower compared with low LRFN4 expression group(P=0.011). Moreover, the link between LRFN4 expression and clinicopathological phases was investigated. (Figure 1C). It was found that LRFN4 expression was closely related to clinicopathological stage. Higher expression level of LRFN4 indicated higher T stage and more regional lymph node metastasis in LUAD(P<0.05). However, LRFN4 expression was not related to M stage(P>0.05). To further verify the expression of LRFN4 in tissues, immunohistochemical staining was performed. From Figure 2, the stained sites were mainly concentrated in the nucleus and the intensity of staining was significantly higher in LUAD tissues than in normal alveolar tissues, which suggested that LRFN4 protein expression is upregulated in LUAD tissues.

**FIGURE 1.**
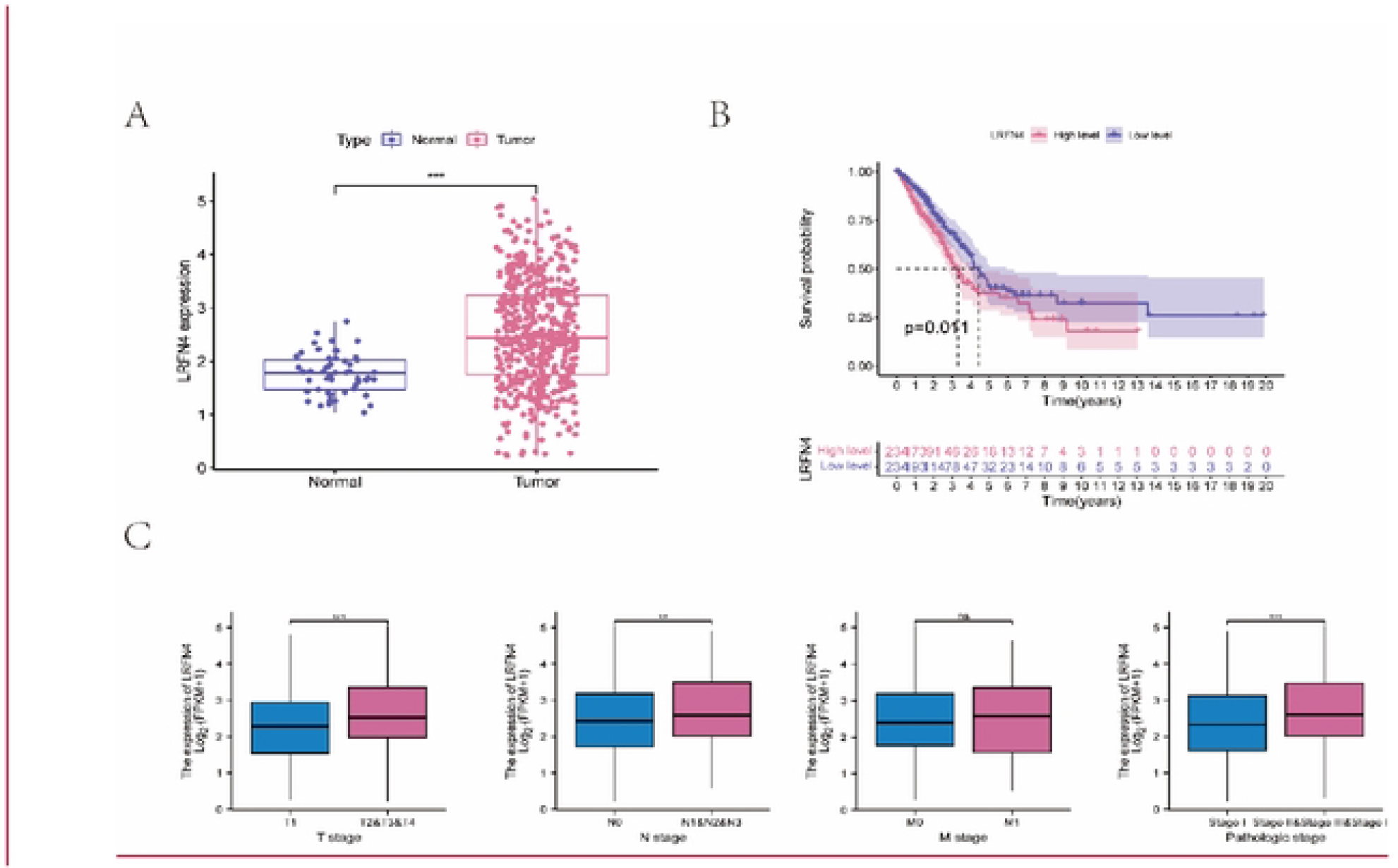
The expression level of mRNA and clinical relevance of LRFN4 :(A) The mRNA expression of LRFN4 in LUAD. (B) Association between LRFN4 expression and survival probability. (C) Association between LRFN4 expression and T sage, N stage, M stage and pathologic stage. (*: P<0.05, **: P<0.01, ***: P<0.001, ns: no significance.)

**FIGURE 2.**
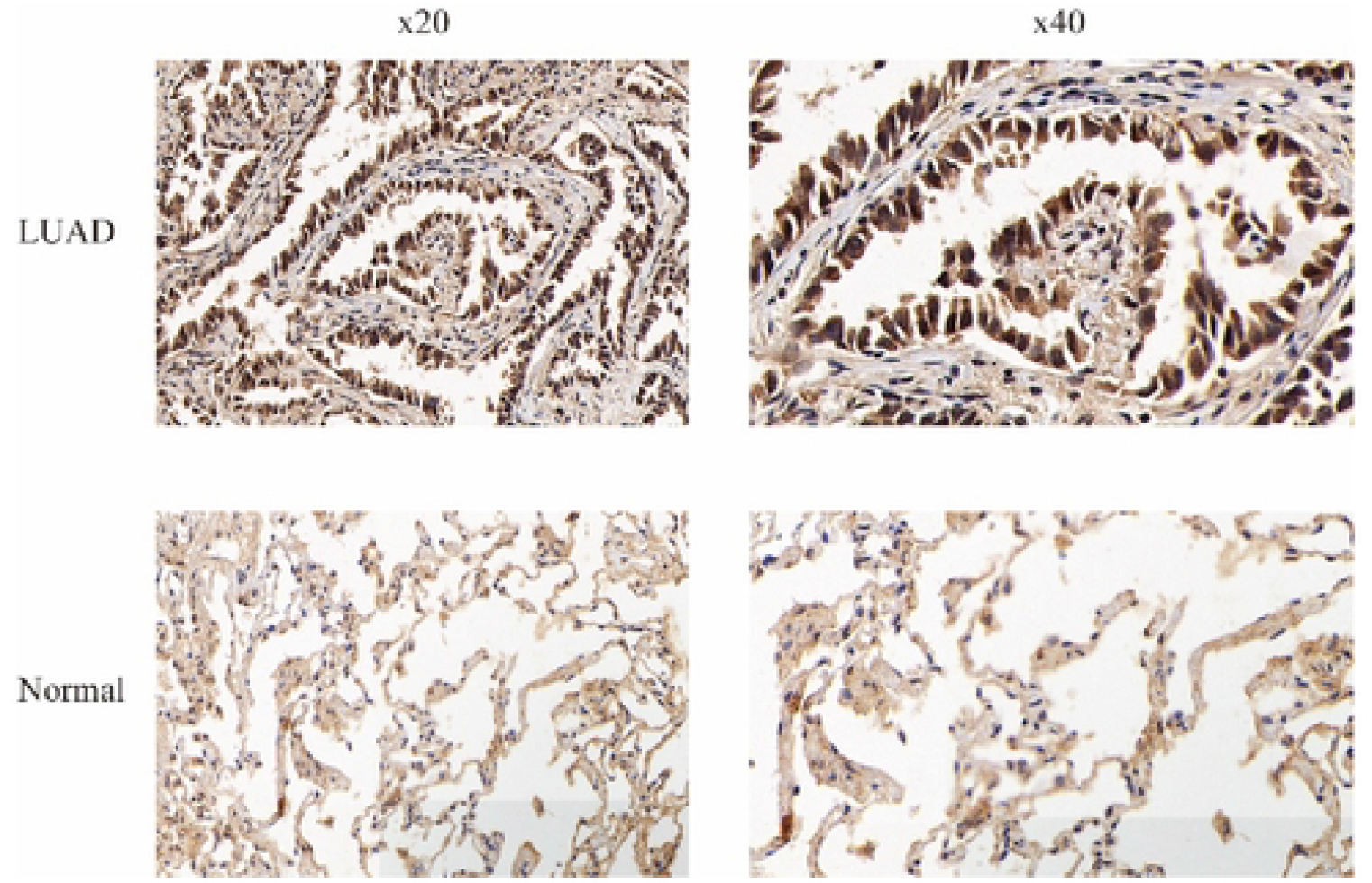
The protein level of LRFN4 in LUAD tissues and normal tissues by immunohistochemical staining.

### LRFN4 expression was related to multiple immune cell infiltration levels

We calculated the contents of 22 immune cells for each LUAD sample and normal sample by CIBERSORT and determined whether they were statistically significant. As shown in figure 3A, there were statistically significant variations in several immune cell infiltration levels between LUAD and normal samples according to our findings, including: naive B cells, plasma cells, CD4+ T cells, regulatory T cells(Tregs), natural killer cells, monocytes, macrophages, dendritic cells, mast cells, eosinophils and neutrophils(P < 0.05). Then, we tried to discover the association between LRFN4 copy number variation and immune cell infiltration levels by TIMER database. With chromosome arm-level gain of LRFN4, infiltration levels of CD4+ T cells (P<0.01) and B cells (P<0.05) were significantly lower (figure 3B). The relationship between more types of immune cells and LRFN4 expression levels was analyzed based on the results of CIBERSORT. The proportions of T cells CD8(P=0.001), T cells CD4 memory activated(P<0.001), T cells follicular helper(P=0.009), macrophages M0(P<0.001) and macrophages M1(P=0.033) were greater in the high LRFN4 expression group than in the low LRFN4 expression group. Conversely, the proportions of T cells CD4 memory resting(P<0.001), T cells gamma delta(P=0.014), monocytes(P<0.001), dendritic cells resting(P<0.001), dendritic cells activated(P=0.003) and mast cells(P<0.001) were lower (figure 3C). Finally, on the TISIDB database, we looked into the relationship between LRFN4 expression and the abundance of various immune cells in LUAD patients. The abundance of a variety of immune cells infiltration was discovered to be linked to LRFN4 expression, including: Th17 cells, NK cells, follicular helper T cells, Act B, effector memory CD4+ T cells, iDC, pDC, imm B cells, Act CD4 and eosinophils. These cells all showed negative correlation with LRFN4 expression, except Act CD4 (figure 4).

**FIGURE 3.**
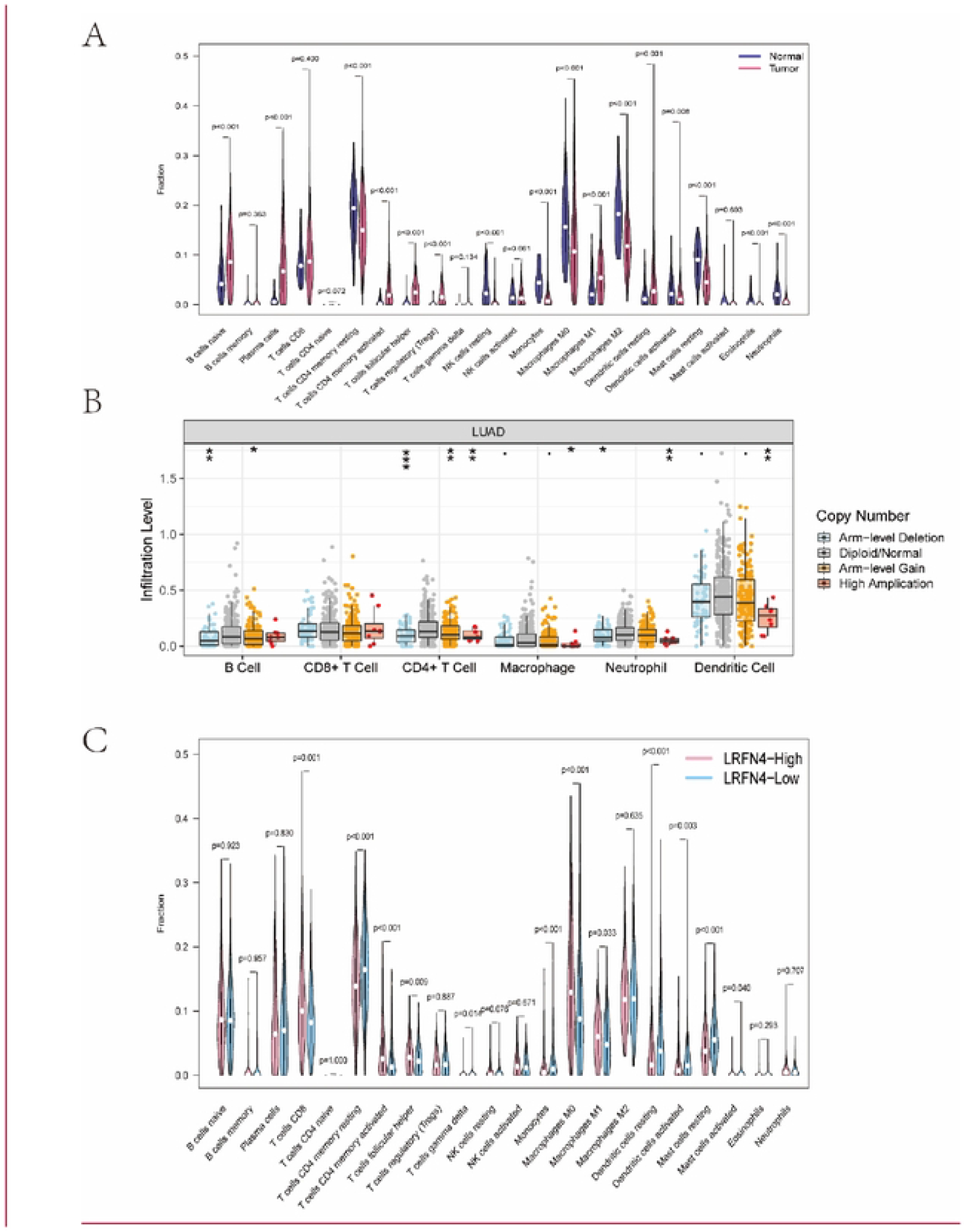
Immune infiltration level in LUAD. (A) 22 immune cell infiltration levels in normal and LUAD tissues. (B) Comparison of several immune cell infiltration levels with different LRFN4 copy number variations in LUAD.(C) Association between the proportions of 22 immune cells and LRFN4 expression in LUAD. (*: P<0.05, **: P<0.01, ***: P<0.001, ns: no significance.)

**FIGURE 4.**
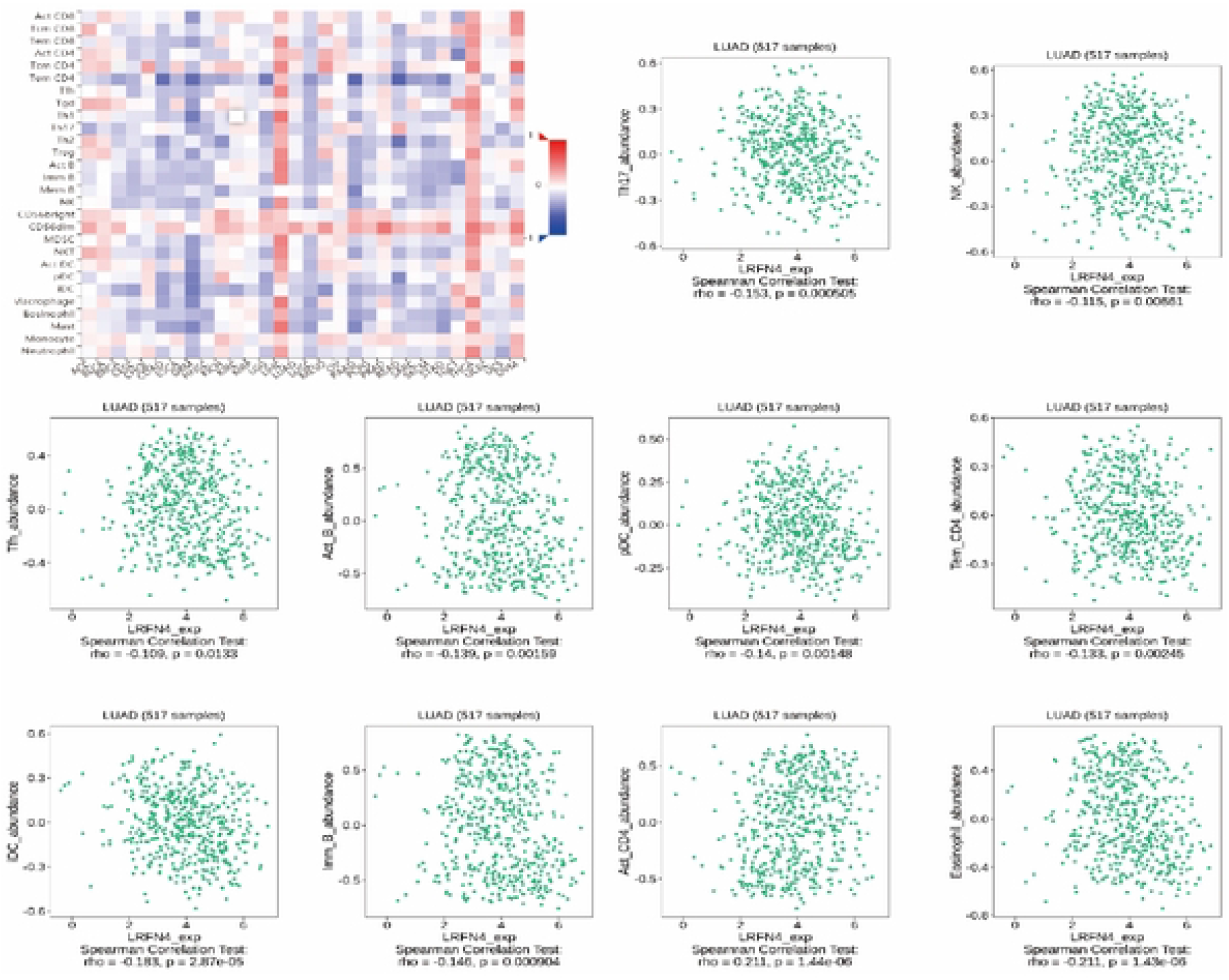
Relationship between the LRFN4 expression and 28 immune cells from TISIDB database.

### Functional enrichment analysis of LRFN4 co-expression molecules and Construction of PPI network

Co-expressed molecules obtained from CCLE database were used for functional enrichment analysis. We set the filtering criteria as co-expression coefficient greater than 0.5 and P-value less than 0.05 and obtained 154 co-expressed molecules of LRFN4 eventually. These compounds were shown to be abundant in 30 GO keywords, including ten biological process terms (BP), ten cellular component terms (CC), and ten molecular function terms (MF) (figure 5A). According to KEGG analysis, they were enriched in five signaling pathways, including: Hippo signaling pathway, inositol phosphate metabolism, phosphatidylinositol system, NOD-like receptor signaling pathway and TNF signaling pathway (figure 5B). From the results of GSEA analysis, we found that several tumor-related biological process and signaling pathways were enriched with high LRFN4 expression: base excision repair, P53 signaling pathway and RNA degradation (figure 5C). We obtained a list of molecules including 8 immunoinhibitors and 27 immunostimulators from TISIDB database(P<0.05). Using the list of molecules, we constructed a PPI network with medium confidence by STRING website (figure 5D).

**FIGURE 5.**
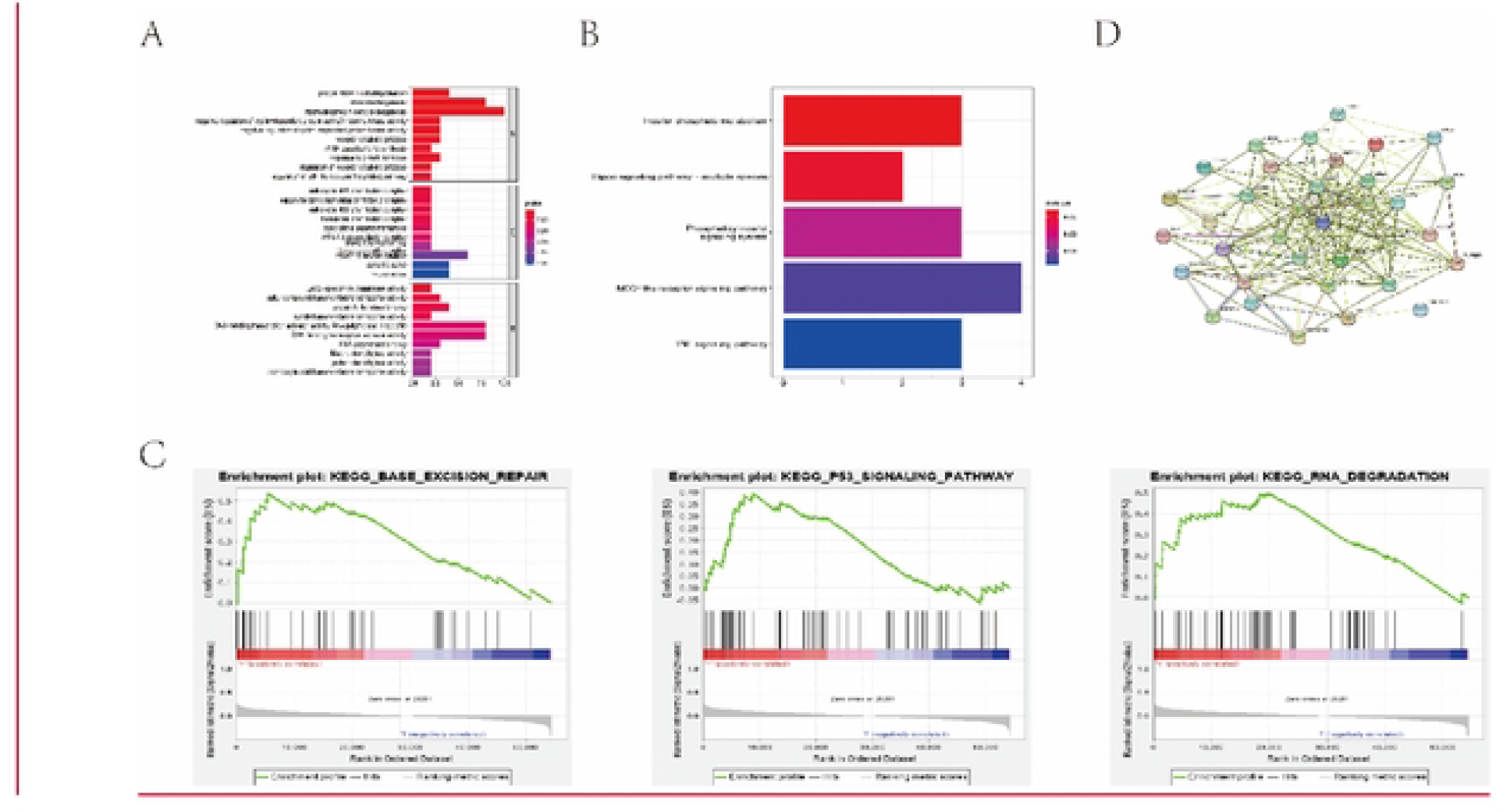
Functional enrichment analysis based on LRFN4-related molecules. (A) Co-expressed molecules of LRFN4 from CCLE database were enriched in 30 GO terms. (B) KEGG pathway analysis of co-expressed molecules of LRFN4. (C) Several tumor-related biological process and signaling pathways were enriched with high LRFN4 expression by GSEA analysis. (D) PPI network of immunomodulators related to LRFN4 from TISIDB database.

### Construction and clinical implications of risk prognostic signature

By univariate COX analysis, we selected 14 immunomodulators that were associated with prognosis in LUAD (p< 0.05). Later, eight immunomodulators (CD40LG, CD48, CD80, CD276, NT5E, PVR, TNFRSF17 and BTLA) were eventually chosen to create a risk prognostic signature by multivariate COX analysis (figure 6A). Among them, CD40LG and CD80 were protective molecules, while CD48, CD276, NT5E, PVR, TNFRSF17 and BTLA were all risk molecules. We can calculate Riskscore for each patient based on the model: Riskscore= (−0.698) *expression level of CD40LG+(0.222)*expression level of CD48+(−0.822) *expression level of CD80+(0.248)*expression level of CD276+(0.191)*expression level of NT5E+(0.191)* expression level of PVR+(−0.191) *expression level of TNFRSF17+(0.732)*expression level of BTLA.

**FIGURE 6.**
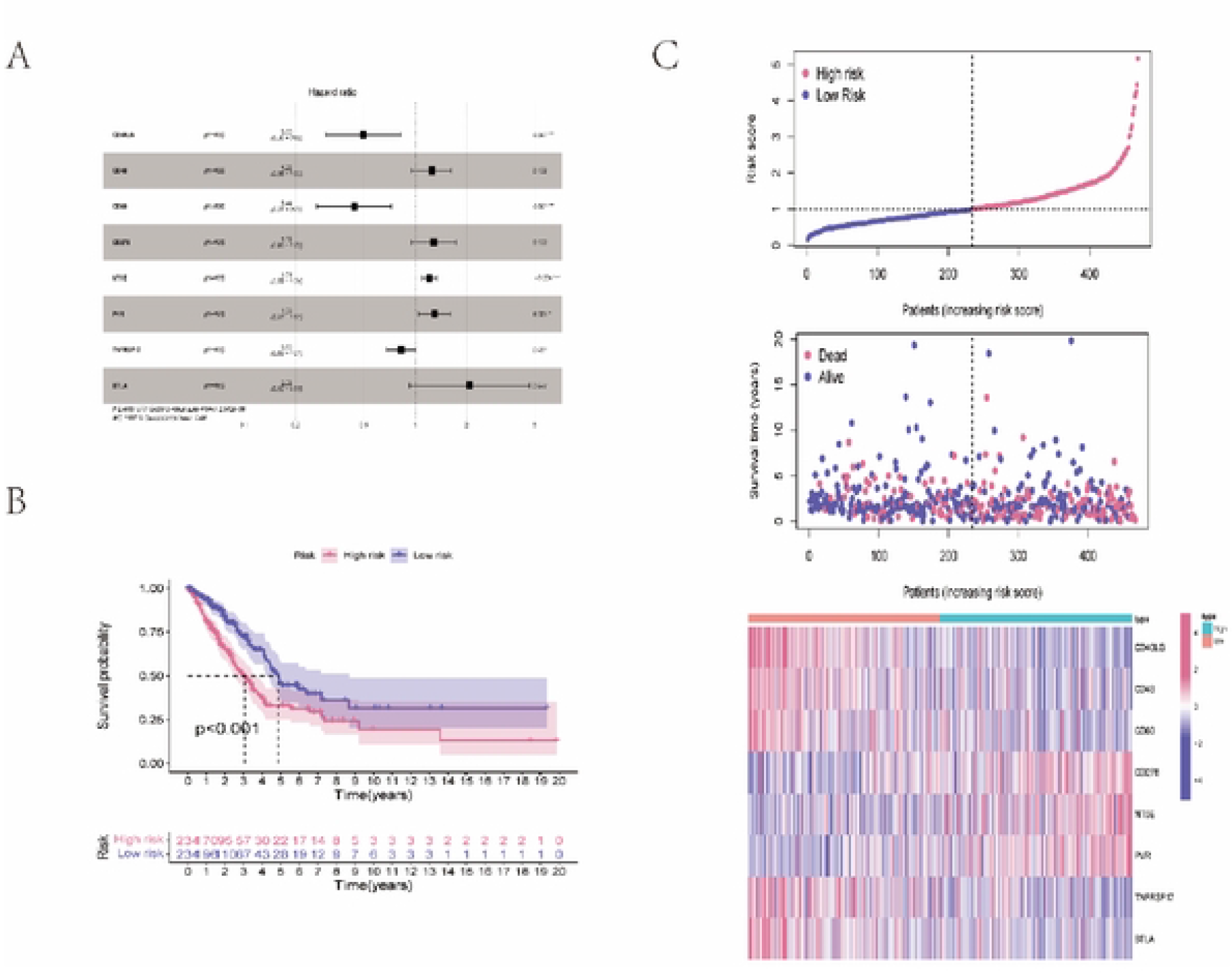
Construction and clinical relevance of risk prediction model (A) Eight immunomodulators associated with prognosis in LUAD by univariate COX analysis. (B) Association between Riskscore and survival probability. (C) Distribution of Riskscores, survival statuses, and gene expression profiles for LUAD.

We performed Kaplan-Meier survival analysis to determine whether Riskscore correlated with clinical outcomes. As shown in figure 6B, Patients with low Riskscores had better prognosis than those with high Riskscores. We also discovered that the number of fatalities was much greater in the high Riskscore category, and the distribution of Riskscores, survival statuses and prognostic signature gene expression profiles were visualized in figure 6C.

We tried to find independent prognostic risk factors for LUAD patients in Riskscore and clinical characteristics (including age, gender and pathological stage). Univariate and multifactorial analysis were performed, as can be seen from figure 7A, pathological stage and Riskscore were independent risk factors for poor prognosis. ROC curve was plotted to examine predictive efficacy, and the area under the curve for Riskscore and pathological stage was 0.653 and 0.687 respectively. However, it can achieve 0.714 with combination of the above two (figure 7B). Finally, a nomogram containing Riskscore and clinical characteristics was constructed, we can predict the probability of survival by calculating total score (figure 7C). We also plotted the calibration curves for 3-year and 5-year which showed that the nomogram’s projected probability (gray line) had a good overlap with the ideal reference line (red line) for LUAD patients. (figure 7D).

**FIGURE 7.**
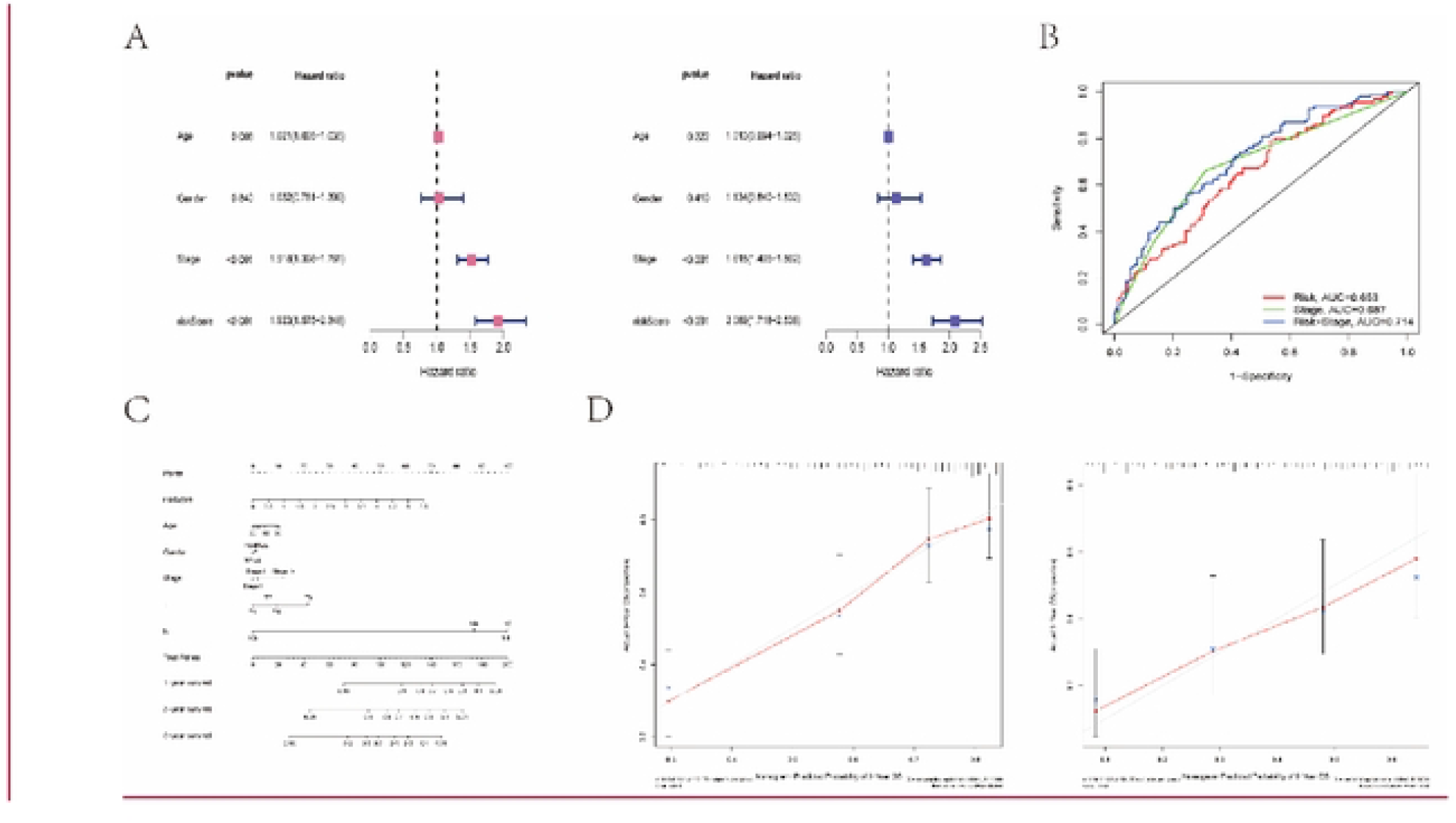
Clinical implications of risk prognostic models (A) Univariate and multivariate COX regression analysis of clinical characteristics and Riskscore in LUAD. (B) ROC curves for clinical characteristics and Riskscore in LUAD. (C) A nomogram for predicting survival possibilities in LUAD. (D) The calibration curves for 3-year and 5-year survival of LUAD patients.

## Discussion

As a neuronal transmembrane protein, the expression and function of LRFN4 were mainly restricted to neural tissue in the past. It was predicted to be involved in regulation of postsynaptic density assembly, presynapse assembly and synaptic membrane adhes ion [20]. In the recent years, it has been reported that LRFN4 is also expressed and function in nonneural tissues. LRFN4 induced monocyte/macrophage migration by reorganizing the actin cytoskeleton, as shown by Shu Konakahara in 2011. Additionally, the researchers observed that LRFN4 was expressed in a wide range of tumors and leukemia cell lines [21]. Songling Han constructed a prognostic model by CTHRC1 and LRFN4 in gastric cancer which showed that low LRFN4 expression was related to poor prognosis for gastric cancer patients [14]. Gang Hu also established a 5-gene model that includes LRFN4 in 2020 and LRFN4 was regarded as a protective gene related to good prognosis in gastric cancer [15]. Nevertheless, according to Ying Liu’s research, elevated LRFN4 expression was linked to a bad prognosis in gastric cancer, which contradicted their findings [13]. F Zheng demonstrated that LRFN4 was a protective gene in colorectal cancer tissue through a retrospective study [17]. Huiqin Li showed that LRFN4 was a risk gene which was related to poor prognosis in ovarian cancer by bioinformatics analysis in 2021[16]. However, the role of LRFN4 in LUAD was still unknown. For the first time, we looked at the link between LRFN4 and LUAD in our research.

Herein, in LUAD, the expression of LRFN4 differed considerably between tumor and para-carcinoma tissues. In LUAD tissues, LRFN4 expression was considerably higher than in normal tissues. This suggested that LRFN4 may act as a biomarker to help us distinguish LUAD tumor tissues from normal tissues. By Kaplan-Meier analysis, we found that when compared to patients with low LRFN4 expression, those with high LRFN4 expression had worse prognosis. Furthermore, high LRFN4 expression was associated with higher T-stage, more regional lymph node metastases and more advanced stage in LUAD, we speculated that LRFN4 may act as a risk gene to induce the proliferation and spread of tumor cells, thus affecting prognosis.

Previous study have reported that tumor microenvironment is vital for the progression of tumors, and influences the patients’ prognosis. B cells, multiple T cells except Tregs, dendritic cells and eosinophils have been shown to favor the survival of LUAD patients [22]. Nevertheless, increased infiltration levels of macrophages M0 and Tregs are associated with poor prognosis [23, 24]. To further verify whether LRFN4 effects the outcomes of LUAD patients by influencing the tumor immune microenvironment, we investigated the relationship between LRFN4 and the level of immune cell infiltration. Herein, we found that the amount of infiltration of various immune cells was linked to LRFN4. LRFN4 expression was negatively correlated with B cell, T helper 17 cell, eosinophil, dendritic cell, follicular helper T cell and NK cell, conversely, LRFN4 expression was positively correlated with macrophages M0. It has been reported that LRFN4 induces transendothelial migration of monocytes/macrophages via actin cytoskeleton reorganization in 2011[21], this suggested that some immune cells’ dispersion may be influenced by LRFN4. The infiltration levels of macrophages M0, macrophages M1, and monocytes were substantially different between the high and low LRFN4 expression groups in LUAD, according to our findings. These findings were consistent with previous studies. We speculated that LRFN4 may reduce the infiltration level of protective cells and increase the infiltration level of risk cells through certain pathways (such as actin cytoskeleton reorganization), thereby affecting patients’ outcomes. As a consequence, LRFN4 may become an immunotherapeutic target that improves the immune microenvironment in LUAD.

The findings of functional enrichment analysis revealed that LRFN4 was linked to a variety of pathways, such as: NOD-like receptor pathway, TNF pathway and Hippo pathway. GSEA analysis results indicated that LRFN4 may be related to base excision repair, P53 signaling pathway and RNA degradation. NOD-like receptors play important roles in the innate immune response. NOD-like receptors mainly include NOD1 and NOD2, and the ligands which they recognize include bacteria and viruses [25]. When bacteria or viruses invade the organism, NOD-like receptors induce immune response by activating multiple pathways, such as: NF-κB pathway, mitogen-activated protein kinase pathway (MAPK pathway), autophagy pathway and endoplasmic reticulum pathway (ER pathway) [26-28]. In addition, NOD-like receptors are also involved in tumorigenesis and development. Da Silva Correia firstly demonstrated the association of NOD1 with tumors. They found that NOD1 acted as a sensitizer of the TNF pathway and promoted apoptosis, and he confirmed this in vitro study using mouse models [29]. Amiri found that chronic activation of NOD1 and NOD2 may be involved in the development and progression of gastric cancer in 2016[30].TNF signaling pathway can induce apoptosis, cell survival or inflammation, the antiapoptotic effect of TNF requires protein synthesis which is mediated by NF-κB activation[31]. Hippo signaling pathway plays an important role in cancer genesis and metastasis, tissue regeneration, and functional regulation of stem cells. An abundance of literature already links dysregulation of the Hippo signaling pathway to cancer progression[32-34].In parallel, P53 signaling pathway serves as a major barrier against the development and progression of tumor. More than half of cancers harbor mutant P53, which inactivates P53 signaling pathway and its tumor suppressor function [35, 36]. In short, LRFN4 may play a non-negligible role in tumorigenesis and progression through multiple pathways. A signature based on immunomodulators related to LRFN4 for predicting LUAD prognosis was constructed. Patients with high Riskscores in the model had lower survival probabilities, which is consistent with our previous Kaplan-Meier analysis. COX univariate and multivariate regression analysis revealed Riskscore as an independent prognostic risk factor in the overall survival rate of LUAD patients. Furthermore, according to ROC curve analysis, we can achieve higher prediction accuracy by combining stage and Riskscore. Later, we established a prognostic nomogram combined Riskscore and clinical characteristics, the fact that the nomogram had a high credibility by validation of calibration curves made it a promising tool for predicting LUAD patients’ outcomes. However, the study has a number of drawbacks. To begin with, our research was based on web databases, when coupled to more experimental validation, the results will be more convincing. Secondly, the role of LRFN4 in various tumor-related pathways need to be demonstrated by deeper study.

According to our findings, LRFN4 expression was dramatically upregulated both at the mRNA and protein levels in LUAD tissues. Meanwhile, high expression of LRFN4 predicted poor prognosis. LRFN4 was associated with multiple immune cell infiltration levels. A signature based on immunomodulators related to LRFN4 achieved good predictive efficacy. LRFN4 was expected to be a diagnostic and prognostic biomarker and LRFN4 may become an immunotherapeutic target for LUAD in the future.

## Data Availability

Date Availability Statement Raw data analyzed in this study was from a public database: TCGA database (https://portal.gdc.cancer.gov/), codes and figures for this study are available from GitHub at https://github.com/Kaifeng306187/Zhiyong-Zhu.

https://portal.gdc.cancer.gov/

## Date Availability Statement

Raw data analyzed in this study was from a public database: TCGA database (https://portal.gdc.cancer.gov/), codes and figures for this study are available from GitHub at https://github.com/Kaifeng306187/Zhiyong-Zhu.

